# Exploring Bottom-Up Visual Processing and Visual Hallucinations in Parkinson’s Disease with Dementia

**DOI:** 10.1101/2020.06.28.20142042

**Authors:** Nicholas Murphy, Alison Killen, Sara Graziadio, Lynn Rochester, Michael Firbank, Mark R Baker, Charlotte Allan, Daniel Collerton, John-Paul Taylor, Prabitha Urwyler

## Abstract

Visual hallucinations (VH) are a common symptom of Parkinson’s disease with dementia (PDD), affecting up to 65% of cases. Integrative models of their etiology posit that a decline in executive control of the visuo-perceptual system is a primary mechanism of VH generation. The role of bottom-up processing in the manifestation of VH in this condition is still not clear. Here we compared amplitude and latency patterns of reversal visual evoked potentials (VEPs) in healthy controls (n=21) and PDD patients (n = 34) with a range of VH severities. PDD patients showed increased N2 latency relative to controls, but patients reporting complex VH (n=17) did not demonstrate any relationship between VEP measurements and their hallucination severity as measured on the neuropsychiatric inventory hallucinations subscale (NPIHal) score. Our VEP findings support previous reports of declining visual system physiology in PDD. However, no notable major relationships between the integrity of the visual pathway and VH were found.

## 1 Introduction

Visual symptoms are common in Parkinson’s disease (PD), and include double vision, dry or painful eyes, poor contrast sensitivity, problems with color vision, and blurring of vision or lowered acuity (Biousse et al., 2004; Davidsdottir et al., 2005; Archibald et al., 2009; Urwyler et al., 2014; Weil et al., 2016). Such problems have been linked to the physical decline of retinal function over the course of disease development with depletion of retinal dopamine (Nguyen-Legros, 1988), and retinal nerve fiber layer thinning (Lee et al., 2014). Electrophysiological measures of visual health, such as the visual evoked potential (VEP), and the electroretinogram (ERG), have been widely used to support the diagnosis of PD as indirect measures of health and integrity of early bottom-up visual processing pathways. Measurements of scalp potentials, as well as scotopic alpha and beta waves generated on the retina during foveal stimulation typically demonstrate a slowing of peak activity in PDD patients relative to controls (Bodis-Wollner and Yahr, 1978; Calzetti et al., 1990; Nowacka et al., 2015), acting as indirect support for pathological evidence of a decline in pre-geniculate visual function (Nguyen-Legros, 1988; Lee et al., 2014).

In 45% of PD cases without dementia (Aarsland et al., 1999; Fénelon et al., 2000), and up to 65% of cases with dementia (PDD) (McKeith et al., 2005), patients will also experience visual hallucinations (VH). The early presence of VH is a strong predictor of cognitive decline (Aarsland et al., 2003), as well as increased mortality and overall reduced quality of life for patients and their carers (Goetz and Stebbins, 1993; 1995). Models of VH in Lewy body dementias (including dementia with Lewy bodies (DLB), and PDD) have posited that VH are a product of the inefficient integration of multiple perceptual sub-divisions of the visual system (Collerton et al., 2005; Shine et al., 2011). The framework for healthy visual perception involves the prediction of sensory inputs expected from the salient features of images (based upon long-term memory of similar images and current context) which are then matched to the actual sensory inputs to minimize any discrepancy between the two.

Thus, perception needs to balance predictions and sensory information. Impairments in cognitive control across executive networks in PDD lead to difficulties balancing these processes, thus impairing the accuracy of matching the visual input to expectations. Despite the precise etiology of VH being unclear, variations in the frequency of visual hallucinations over the course of disease progression suggests that these hallucinations reflect a complex relationship between declining sensory function and dysfunctional predictions (Collerton et al., 2005; Onofrj et al., 2007; Fenelon, 2008; Llebaria et al., 2010; Sanchez-Castaneda et al., 2010; Shine et al., 2011).

In this investigation we sought to characterize the components of early bottom-up processing in PDD patients, using the pattern reversal visual evoked potential, and to relate the response features to the complexity of the VHs experienced. Based on available evidence of physiological decline in PDD we predicted that we would observe a general reduction in the amplitude of the VEP components, as well as an increase in the P1 latency (Matsui et al., 2005). In addition we expected baseline visual acuity and visual perception, to demonstrate a decline in those with a more severe and frequent complex VH. This should also extend to an association between VEP P1 and N2 measurements with VH experience, as both of these are thought to be contingent upon attentional and perceptual processes (Haider et al., 1964; Luck 2005), which are, in particular, disrupted by Lewy body pathology (Shine et al., 2011; Taylor et al., 2011).

## 2 Methods

### 2.1 Participants

A total of 21 healthy controls, and 38 Parkinson’s disease with dementia (PDD) patients were recruited from the North East of England. Ethical approval was granted by the Newcastle National Health Service (NHS) Health Research Authority (HRA) (REC reference: 13/NE/0252; R&D reference: 6691). The diagnosis of PDD was confirmed by two independent and experienced clinicians (Charlotte Allan, John-Paul Taylor) and met with the standards described in the international PD diagnostic criteria (Emre et al., 2007). Participants were excluded from the study if baseline assessment revealed the presence of comorbid factors including stroke, non-PD related dementia, and/or visual dysfunction secondary to glaucoma. All procedures related to the study were explained to the participants and written informed consent was obtained prior to participation.

### 2.2 Clinical assessments

All participants were assessed on their level of global cognitive function using the Mini Mental State Exam, (MMSE, (Folstein et al., 1975); maximum score of 30) and the Cambridge Cognitive Test Battery (CAMCOG total score, (Roth et al., 1986; Roth et al., 1988); maximum score of 107). Motor function was assessed using the total (left and right) score from the Unified Parkinson’s disease rating scale section three (UPDRS-III, (Fahn et al. 1987); maximum score of 57).

The integrity of the participant’s visual acuity was assessed using a detailed screening questionnaire, computerized Freiburg acuity testing (Bach, 1996), and the LOGMAR (Logarithm of the Minimum Angle of Resolution) scale of visual acuity. Visuo-perceptual function was assessed using performance on motion sensitivity, (Wood et al., 2013), angle discrimination (Wood et al., 2013), and performance on the pareidolic imagery test (Uchiyama et al., 2012).

### 2.3 Visual Hallucinations

The hallucination subscale of the Neuropsychiatric Inventory (NPIHal) (Cummings et al., 1994) was used for assessing VH occurring in the previous month, with the NPIHal score (frequency × severity of hallucinations) derived as a measure. For reliability, patients and carers were independently asked about the occurrence of VH in the month before using the North-East Visual Hallucinations Interview (NEVHI) (Mosimann et al., 2008). Any discrepancies in the reporting of VH (Urwyler et al., 2015) were discussed with both parties and the assessor, with reformulation of NPIHal scores (wherever the patient seemed to lack insight, primacy was given to caregiver opinion).

Participants were classed as active visual hallucinators (PD-VH, n=17) if they had complex VH in the month preceding their interview; otherwise, they were classed as non-hallucinators (controls (n=21) and PD-NVH(n=17)). Participants with minor VH (e.g., passage or feeling of presence) but no complex VH in the last month were included in the PD-NVH group. This distinction was made due to the different etiologic basis to complex VH even though minor VH typically precede complex VH. Patients in this study map onto the same categories used in previously published research from our lab (see Firbank et al., 2018).

### 2.4 EEG

#### 2.4.1 Visual Evoked Potential Presentation and Recording

The VEP adhered to the specifications proposed by the International Society for Clinical Electrophysiology of Vision (Odom et al., 2010) (ISCEV). Participants viewed a black and white checkerboard pattern whilst the checks (visual angle of 0.6°) reversed phase at a rate of 1Hz (switching to the opposite phase every 500ms), for 200 sweeps, with a brief rest period (3000ms) after 100 sweeps. During stimulus presentation a pink dot was placed in the center of the display as a focus point, which the participant was instructed to look at. This was intended to prevent wandering gaze during the check reversal and was presented on top of a grey background during the rest period. The stimulus was generated on a Dell OptiPlex 755 (Microsoft Windows XP) using Matlab v2012a (The MathWorks, 2012), and presented using a Dell U2412M 24-inch LCD monitor (resolution: 1920 × 1200 pixels refresh rate: 60Hz). Pattern reversal visual evoked potentials were recorded during three separate viewing conditions (both eyes, left eye, right eye), using an ASA-LAB 136 system amplifier and the ASA-LAB recording software (version 4.9.1) in combination with a 128 Ag/AgCl channel Waveguard cap (10-5 system, (Oostenveld and Praamstra, 2001) Advanced Neuro Technologies). The ground electrode was placed on the right clavicle, and Fz was used as the reference electrode. Electrode impedance was kept below 5kΩ, and no filters were applied during the acquisition of EEG data.

#### 2.4.2 Pre-Processing

Signal processing and measurement was performed using Matlab v2012a (The MathWorks, 2012), with the EEGLab (Delorme and Makeig, 2004), ERPLab (Lopez-Calderon and Luck, 2014), and current source density (Kayser and Tenke, 2006) (CSD) toolboxes. Individual sweeps were split into epochs of 400ms, a baseline period of 100ms prior to stimulus presentation, and a post-stimulus period of 300ms. Epochs were baseline corrected using the mean of the data in the pre-stimulus period and filtered using a 0.1 to 45Hz bandpass filter. Individual channels with a kurtosis value greater than three standard deviations from the cap-wide mean were removed and recreated after pre-processing using spherical interpolation (Perrin et al., 1989; Ferree, 2000; Delorme and Makeig, 2004; Ferree, 2006). After removing trials containing blinks, muscular activity, and drifting potentials (impedance related artefacts), broad spatial effects of the electric field were attenuated by applying a Laplacian transform (Kayser and Tenke, 2006). This approach was applied to reduce the likelihood of false positives in spatially distant locations when defining the occipital region of interest (ROI).

#### 2.4.3 Measurement

To account for individual variance in the timing of synaptic communication the VEP components were measured within windows defined by the global field power (GFP) for each individual. The VEP components elicited three GFP maxima following stimulus presentation, each of which was used as the center point for the corresponding component window (GFP maxima ±10ms). The occipital ROI was defined by measuring the amplitudes of the P1 component for the grand average of the control data set and using the 20 electrodes with the greatest amplitude as the limit for the ROI. Individual subject measurements of peak latency and mean amplitude were taken from the average VEP waveform within the occipital ROI. To account for potential inter-ocular latency differences we estimated the difference between P1 peak latency measurements for the left and right eyes.

#### 2.4.4 Statistical Analysis

All statistical tests were performed using the Statistical Package for the Social Sciences (SPSS, version 22). Demographic and baseline factors were compared using independent samples t-tests. We compared the measurements of amplitude and latency separately for each component using univariate analysis of variance controlling for age and inter-ocular latency difference between the left and right eyes. Effect sizes were estimated using the partial eta squared measure (η2). To explore the relationship between the variance within our physiological measurements and VH experience in the hallucinating PDD group, we performed Spearman’s correlations between each VEP measurement and NPI hallucination subset score. To help identify any variance in our measurements accounted for by clinical and/or demographic factors we performed additional Spearman’s correlations between the VEP measurements and each value. Significance for all tests was determined using an alpha criterion of p<0.05, and Bonferroni corrected for multiple comparisons (corrected alpha criterion of p<0.016). Where appropriate un-corrected correlations are reported to highlight trends within individual results.

## 3 Results

### 3.1 Demographics & Clinical Scores

Demographic results are summarized in Table 1. All groups were matched for age and there were no significant differences in duration of PD or levodopa dose between the PDD-VH and PDD-NVH groups. PDD patients displayed a significant reduction in global cognitive function, UPDRS motor score relative to controls, with the PDD-VH group global cognitive function and motor function were significantly worse when compared to the PDD-NVH group.

**Table 1.** Participant demographics and clinical scores. *Denotes significant at *p*<0.05.

| Measurement | Controls<br>(n=21) | PDD NVH<br>(n=17) | PDD VH<br>(n=17) | Posthoc | Statistics<br>*Test Val, p, Effect Size* |
| --- | --- | --- | --- | --- | --- |
| Age (years) | 74.90 ± 5.16 | 72.94 ± 5.19 | 73.88 ± 5.36 |  | 0.756, 0.524, 0.026 |
| MMSE score* | 29.10 ± 1.81 | 24.59 ± 5.0 | 22.76 ± 4.99 | HC>VH, HC>NVH | 12.307, <0.001, 0.321 |
| CAMCOG total score* | 95.14 ± 6.79 | 80.94 ± 15.53 | 73.18 ± 15.91 | HC>VH, HC>NVH,<br>VH<NVH | 14.003, <0.001, 0.350 |
| CAMCOG Memory score* | 23.52 ± 1.5 | 19.94 ± 4.93 | 17 ± 4.1 | HC>VH, HC>NVH | 15.32, <0.001, .36 |
| CAMCOG Executive score* | 22.28 ± 3.16 | 14.53 ± 3.93 | 12.44 ± 4.04 | HC>VH, HC>NVH | 38.67, <.001, .59 |
| UPDRS III score* | 2.10 ± 2.47 | 38.65 ± 21.93 | 57.88 ± 20.47 | HC<VH, HC<NVH,<br>VH>NVH | 55.147, <0.001, 0.680 |
| Acuity (decimal)* | 0.58 ± 0.31 | 0.29 ± 0.21 | 0.31 ± 0.19 | HC>VH, HC>NVH | 7.783, 0.01, 0.234 |
| Acuity (logmar)* | 0.31 ± 0.26 | 0.65 ± 0.32 | 0.57 ± 0.27 | HC>VH, HC>NVH | 7.823, 0.001, 0.235 |
| Minimum Angle Perception<br>(degrees)* | 8.63 ± 3.25 | 28.42 ± 23.51 | 32.68 ± 30.07 | HC<VH | 7.059, 0.002, 0.214 |
| Motion Perception* | -2.72 ± 0.72 | 1.80 ± 3.15 | 2.68 ± 2.88 | HC<VH | 26.746, <0.001, 0.522 |
| Number of Pareidolia* | 1.0 ± 1.46 | 3.18 ± 4.54 | 6.82 ± 5.58 | HC<VH, VH>NVH | 8.188, 0.001, 0.254 |
| Levodopa Dose (24 hours, mg) |  | 569.12 ± 303.05 | 710.59 ± 363.10 |  | -1.04, 0.31, 0.35 |
| Age at Onset of PD symptoms<br>(years) |  | 64.65 ± 7.08 | 60.88 ± 7.62 |  | 1.06, 0.29, 0.36 |
| PD Duration (years) |  | 7.18 ± 4.51 | 10.82 ± 7.46 |  | -1.47, 0.15, 0.5 |
| NPI total score* |  | 0.29 ± 0.58 | 3.11 ± 2.05 | NVH<VH | -5.44, <.001, 3.23 |

Data are mean ± standard deviation. Statistical tests: : Univariate analysis of variance (ANOVA), p-value 2 sided <0.05 significant; PD = Parkinson’s disease, PDD = Parkinson’s disease dementia, MMSE = Mini-Mental State Examination, CAMCOG = Cambridge Cognitive Assessment, UPDRS = Unified Parkinson’s Disease Rating Scale, HC = Healthy Controls, VH = Visual Hallucination, NVH= No Visual Hallucination; Tests reported used univariate ANOVA with partial eta squared effect size, except for Levodopa, age at onset, duration of PD, and NPI Hal total which used independent samples t-tests and Cohen’s D effect sizes.

### 3.2 Visual Integrity & Visual Perceptual Scores

Visual acuity and perceptual scores are summarized in Table 1. There was a pattern of overall decline in visual integrity within the PDD patients relative to the control group, characterized by a significant reduction in decimal and LOGMAR measurements of visual acuity. As expected, PDD-VH patients showed a characteristic significant increase in the number of false perceptions reported during the pareidolia task compared to PDD-NVH patients.

### 3.3 Visual Evoked Potential

Amplitude tended to be smaller, and latency later in PDD vs controls, although this was not significant, except for N2. There were no significant differences between PDD-VH vs PDD-NVH. (see Table 2). Follow up simple effects analysis demonstrated that N2 latency in controls was significantly less than PDD-VH (p = .022) and PDD-NVH group (p = .03), but the N2 latency did not differ between the VH and NVH group.

**Table 2.** Comparison of the visual evoked potential component (N1, P1 and N2) amplitude and latency.

| Component | | Controls<br>(n=21) | PDD NVH<br>(n=17) | PDD VH<br>(n=17) | Statistics<br>F, p-value, $\eta^2$ |
| --- | --- | --- | --- | --- | --- |
| **N1** | Amplitude | -1.27 ± 0.93 | -0.85 ± 0.82 | -0.84 ± 0.51 | 1.63, 0.21, 0.06 |
| | Latency | 88.28 ± 8.62 | 90.23 ± 08.68 | 93.35 ± 10.47 | 0.46, 0.63, 0.02 |
| **P1** | Amplitude | 3.61 ± 2.55 | 2.32 ± 1.82 | 2.36 ± 1.33 | 2.16, 0.13, 0.08 |
| | Latency | 124.50 ± | 127.59 ± 7.73 | 126.84 ± 6.33 | 1.47, 0.24, 0.06 |
| **N2** | Amplitude | -1.64 ± 1.44 | -0.97 ± 0.84 | -1.26 ± 1.19 | 1.7, 0.19, 0.06 |
| | Latency<br>(ms) | 162.27 ±<br>8.97 | 176.93 ±<br>14.69 | 174.15 ±<br>15.30 | 7.44, **0.001\***, 0.23 |

Data are mean ± standard deviation. Statistical tests: Univariate analysis of variance (ANOVA), $df=52$ , $p$ -value 2 sided $<0.05$ significant; PDD = Parkinson's disease dementia; HC = Healthy Controls; VH = Visual Hallucination; NVH= No Visual Hallucination; \* Posthoc = VH>HC, NVH>HC;

### 3.4 Clinical Correlations

Visual hallucination experience, as measured using the NPIHal subscale was not significantly related to the measurements of any of the VEP components. In PDD-VH patients there were no significant correlations between any of the VEP measurements, demographic, and clinical factors.

## 4 Discussion

In healthy participants, the VEP reflects a combination of many pre-striate and cortical processes. It is noted that a decline in visual pathway integrity following structural changes to the retina can affect the latency and amplitude (Bodis-Wollner and Onofrj, 1982; Bhaskar et al., 1986; Nowacka et al., 2015; Miri et al., 2016). In earlier studies the VEP has consistently been shown to be affected by PD neuropathology, indicating substantial decline in the quality of bottom-up visual processing (Archibald et al., 2009; Bodis-Wollner & Yahr, 1978; Nowacka et al., 2015). Following the hypothesis that disrupted bottom-up processing of visual input is associated with the generation of VH in PDD we investigated whether the VEP could be used as a marker of hallucination symptomology.

In accordance with previous research (Mosimann et al., 2004; Emre et al., 2007; Archibald et al., 2009; Possin, 2010; Landy et al., 2015) the PDD patients demonstrated a reduction in visual acuity, impaired visual perception, impoverished motor ability, and compromised global cognition. Analysis of the pattern reversal VEP data revealed a significant increase in the PDD N2 latency relative to controls, especially in PDD-VH, and non-significant reduction in the PDD P1 amplitude. P1 and N2 (N140) are both linked to physical properties of the stimulus such as luminance, brightness, position on the retina, and associated attentional demands (Van Voorhis and Hillyard, 1977; Hillyard and Munte, 1984; Luck et al., 1994; Johannes et al., 1995; Ito and Gilbert, 1999; Johannes et al., 2003). Further, the N2 (N140) has been reported to be associated with increased disease severity (Talebi et al, 2014). In patients with PDD there are often abnormalities associated with the structure and function of the retina, including changes in morphology and dopaminergic signaling (Archibald et al., 2009), which have previously been linked to reduced conduction velocity in early visual processing (Regan and Neima, 1984; Bodis-Wollner et al., 1987; Jones et al., 1992; Price et al., 1992; Pieri et al., 2000; Holroyd et al., 2001; Nowacka et al., 2015). Source localization of these components places the generating sources deep within the secondary visual cortex (Di Russo et al., 2002; Di Russo et al., 2005); although their cognitive associations suggest that their activity is governed as part of a higher order visual processing network. Given the lack of association between the VEP components and clinical measurements in our study it is unclear what relationship exists between the primary visual cortex and its bottom-up and top-down inputs in this context. However, our experimental design is limited in the scope to which we can draw conclusions on the nature of pathological change within the executive system and the link between attention and passive perception of the VEP stimulus.

In the context of a mechanism for VH, our sample results suggest that bottom-up processing is not differentially affected between hallucinators and non-hallucinators. This is not unexpected as it follows that in an integrative model of VH we would expect VH content to stem from the interaction of impaired bottom-up processing with dysfunctional top-down control of perception. In our data, complex VH were associated with greater decline in CAMCOG, and UPDRS scores, as well as increased numbers of pareidolia relative to patients without complex VH. The divergence in the cognitive and perceptual profile of the groups supports a deteriorated capacity for effective top-down control, which in this model would be a pre-requisite factor for the generation of complex VH. However, these measures were not significantly correlated with the amplitude or latency of the VEP component measurements suggesting that conduction velocity and basic processing of visual feature information is unimpeded by the integrity of detailed perceptual processing.

Within the integrative model of complex VH in Lewy body dementias the importance of bottom-up processing is thought to be its influence on the generation of proto-objects (Collerton et al., 2005; Shine et al., 2011). The frequency and phenomenology of the VH would then depend on the interaction between the executive system and the perceptual processing centers. Therefore, declining visual health and perceptual quality might simply place the individual in an at-risk state for VH development (Firbank et al., 2018) rather than directly impact their generation. Further research is required to model the way pathological effects on top-down processing interact with declining visual health.

### 4.1 Limitations

There are several limitations. Firstly, the sample size within this study was relatively small. Secondly, we used only a single subjective measure for VH severity. The NPI items are typically collected from the carers of the patient, and do not ask questions about the content of the hallucination. It thus remains possible that there could be a relationship between visual health, bottom-up processing, and VH content that could be accessed by quantifying a scale such as the North East Visual Hallucination Interview (NEVHI) (Mosimann et al., 2008). Furthermore, the range of VH severity scores in our groups was limited making correlative analyses more difficult.

### 4.2 Conclusion

In summary, PDD patients demonstrated a diminished profile for visual information processing by way of lowered acuity and reduced visual integrity. This was partially reflected in the outcome of the VEP components, although the broad lack of significant differences between PDD-VH, PDD-NVH, and healthy controls implies that bottom-up visual information processing remains reasonably intact. Our findings support a separation between bottom-up information processing and the mechanism of complex VH generation, and instead imply that the reduced visual integrity might act to place the individual in an at risk state for the development of hallucinations in patients with a deteriorated cognitive profile. Future work should focus on a multimodal approach to understanding the interactions between top-down and bottom-up perceptual circuitry and how this is impacted by PDD neuropathology.

## Data Availability

Data pertaining to the obtained results may be provided upon request.

## 6 Author contributions

NM, SG, LR, CA, DC, and J-PT contributed to the conception and organization of the research. NM and AK participated in the execution and data collection. NM, PU and MF designed and implemented the data analysis and interpretation. NM and PU wrote the first draft of the manuscript. All authors approved the final version of this manuscript.

## 8 Acknowledgements

We would like to thank all participants for taking part. The authors are grateful to the group of Professor Etsuro Mori, Tohoku University School of Medicine, Sendai, Japan, for providing a copy of the pareidolia task.

## 9 Data Availability

Data pertaining to the obtained results may be provided upon request.

## 10 Figure Captions

Table 1 | Participant demographics and clinical scores. Data are mean ± standard deviation. Statistical tests: : Univariate analysis of variance (ANOVA), p–value 2 sided <0.05 significant; PD = Parkinson’s disease, PDD = Parkinson’s disease dementia, MMSE = Mini-Mental State Examination, CAMCOG = Cambridge Cognitive Assessment, UPDRS = Unified Parkinson’s Disease Rating Scale, HC = Healthy Controls, VH = Visual Hallucination, NVH= No Visual Hallucination; Tests reported used univariate ANOVA with partial eta squared effect size, except for Levodopa, age at onset, duration of PD, and NPI Hal total which used independent samples t-tests and Cohen’s D effect sizes.

Table 2 | Comparison of the visual evoked potential component (N1, P1 and N2) amplitude and latency. Data are mean ± standard deviation. Statistical tests: Univariate analysis of variance (ANOVA), df=52, p–value 2 sided <0.05 significant; PDD = Parkinson’s disease dementia; HC = Healthy Controls; VH = Visual Hallucination; NVH= No Visual Hallucination; * Posthoc = VH>HC, NVH>HC;

